# The IGNITE Trial: Participant Recruitment Lessons Prior to SARS-CoV-2

**DOI:** 10.1101/2020.05.19.20107458

**Authors:** Eric D. Vidoni, Amanda Szabo-Reed, Chaeryon Kang, Jaime Perales-Puchalt, Ashley R. Shaw, George Grove, Morgan Hamill, Donovan Henry, Jeffrey M. Burns, Charles Hillman, Arthur F. Kramer, Edward McAuley, Kirk I. Erickson

## Abstract

Full and diverse participant enrollment is critical to the success and generalizability of all large-scale Phase III trials. Recruitment of sufficient participants is among the most significant challenges for many studies. The novel SARS-CoV-2 coronavirus pandemic has further changed and challenged the landscape for clinical trial execution, including screening and randomization. The Investigating Gains in Neurocognition in an Intervention Trial of Exercise (IGNITE) study has been designed as the most comprehensive test of aerobic exercise effects on cognition and brain health. Here we assess recruitment into IGNITE prior to the increased infection rates in the United States, and examine new challenges and opportunities for recruitment with a goal of informing the remaining required recruitment as infection containment procedures are lifted. The results may assist the design and implementation of recruitment for future exercise studies, and outline opportunities for study design that are flexible in the face of emerging threats.

## INTRODUCTION

Many countries around the world, including the United States, are experiencing a rapid demographic shift, with greater numbers of individuals living longer.[1] Associated with increased life expectancy is an increase in health care costs and risk of chronic and debilitating age-related diseases such as Alzheimer’s disease (AD). The rising number of individuals at risk for cognitive decline is accelerating efforts to develop effective treatment and prevention approaches as policy makers, clinical researchers, drug developers, and other stakeholders fully recognize the scope of the problem. Exercise is a promising approach for improving brain and cognitive health in late adulthood [2-5] but the field still lacks definitive evidence of benefit.[6] Our Phase III multi-site randomized dose-response clinical trial called IGNITE (NCT02875301) has been designed to definitively address whether exercise impacts cognitive and brain health in cognitively normal older adults.[7]

Clinically definitive trials generally require large sample sizes (e.g., >500) to evaluate the primary outcome of interest. Efficiently and effectively recruiting adequate numbers of qualified volunteer participants is among the biggest challenges facing cognitive aging investigators.[8] Nearly 20% of trials fail to adequately recruit the number of participants necessary to address their clinical question.[9] Trial recruitment is commonly cited as being among the most costly barriers to advancing our understanding of cognitive health interventions[8, 10, 11] and requires significant investment of money, institutional resources, time, and personnel.[12] The pace of recruitment into trials directly impacts the cost of trials and their time to completion.[10, 13] Additionally, for the results of such trials, including IGNITE, to be truly generalizable to the broader aging population, the sample should strive for participant heterogeneity, including ethnic, racial, sex, socioeconomic, and geographic representation. Long-standing exclusion and exploitation of underrepresented individuals, whether due to racism, sexism, disenfranchisement, or distance from scientific or university facilities, increases the importance, but also the difficulty of heterogeneous recruitment.[14, 15]

In late December 2019, the novel coronavirus known as SARS-CoV-2 (severe acute respiratory syndrome coronavirus 2) was first reported. By late February of 2020, United States municipalities began applying a variety of infection containment policies such as social distancing and non-essential business closures to control the spread of SARS-CoV-2. These policies significantly impacted clinical research activities and recruitment into most clinical research across the United States,[16] including IGNITE. All IGNITE sites temporarily halted enrollment and in-person study activities consistent with institutional guidance. Recognizing that recruitment and study procedures would likely be altered for the remainder of the trial timeline, the IGNITE team undertook an assessment of trial recruitment up to the SARS-CoV-2 closures, to assess (a) lessons learned regarding multi-site exercise trial recruitment with respect to demographics (especially underrepresented ethno-racial minorities, URM) and study procedures, and (b) potential new recruitment barriers and opportunities after containment policies were eased. The goal of the analysis was descriptive in nature, rather than driven by *a priori* hypotheses. Expected outcomes were identified barriers and opportunities to enrollment post SARS-CoV-2 onset applicable to large-scale exercise trials for healthy, sedentary older adults.

## METHODS

IGNITE was designed to address several important unanswered questions: (1) Are the recommended public health guidelines of 150 min/week of moderate intensity exercise sufficient for improving cognitive performance? (2) At that dose, does exercise influence brain structure and/or function? (3) Is there a dose-response effect of exercise on cognitive performance or brain structure/function, such that exercise that exceeds the recommended levels of 150 min/week results in even greater benefits in cognitive and brain health? (4) Can we identify possible mechanisms (e.g., cardiometabolic, inflammatory, neurotrophic, or psychosocial changes) by which exercise influences cognitive and brain health? (5) Are there factors (e.g., demographic characteristics, presence of brain beta amyloid, genotype) that attenuate or magnify the effects of exercise on brain, cognitive, and psychosocial health and contribute to the individual variability in intervention outcomes? And, (6) could individual differences or changes in beta amyloid accumulation as a function of participation in exercise explain any cognitive, brain, or psychosocial improvements?

The IGNITE protocol has been detailed previously.[7] Briefly, IGNITE participants are administered a two-part cognitive and psychosocial test battery, graded maximal exercise treadmill test, physical function battery, MRI/fMRI, florbetaben PET scan, dual-energy x-ray absorptiometry scan, blood and hair collection at multiple time points over one year. Physical activity is assessed bi-monthly by accelerometry. Enrollees passing all screening criteria are randomized to one of three groups: 150 min per week of light intensity stretching and toning condition exercise; 150 min per week of aerobic exercise; 225 min per week of aerobic exercise. Recruitment, enrollment, and randomization occurs on a rolling basis.

We set recruitment goals for racial and ethnic minorities in proportion to the demographic representation surrounding each of the three study sites: Northeastern University, Boston, MA (25% black; 14% Hispanic; 7.3% Asian); University of Kansas Medical Center, Kansas City, KS-MO (11% black; 2.8% Hispanic; 1.5% Asian); University of Pittsburgh, Pittsburgh, PA (26% black; 2.3% Hispanic; 4.4% Asian). We anticipated approximately 60% of the final sample would be female.

Each study site used recruitment strategies specific to the local resources and environments as well as prior experiences of successful recruitment strategies in other previous studies although many of the recruitment strategies were similar across sites (described below). Planned recruitment approaches included both proactive (e.g. calls to participants in health system research registries, presentations at senior centers and in faith communities) and passive strategies (newspapers, health direct mailings, and social media). Phone screenings were conducted by each site through a central database (REDCap[17]) custom designed by the University of Pittsburgh coordinating center. The phone screening form contained standard screening language employed by all sites. Inclusion and exclusion criteria can be found in Table 1. The script began with a description of the study design, and then study procedures, and request for verbal consent for data collection to determine eligibility. This was followed by demographic information collection, eligibility criteria confirmation most likely to exclude an individual, and finally the Telephone Inventory of Cognitive Status.[18] The phone screen was built with dependency logic that was self-terminating, such that a phone screen would end if any exclusionary criteria were identified at any point, or if the participant declined for any reason, prior to completing the full screen. Therefore, not all demographic, inclusion or exclusion criteria, or medical conditions were captured for each potential participant, resulting in a sparse data set. Free text notes were made regarding participant or staffing inclusion concerns and were manually coded when available.

**Table 1.** Inclusion and Exclusion Criteria

| Inclusion criteria | Exclusion criteria |
| --- | --- |
| • Age 65–80 yrs.<br>• Ambulatory without pain or the use of assisted walking devices<br>• Able to speak and read English<br>• Medical clearance by primary care physician (PCP)<br>• Living in community for duration of the study<br>• Reliable means of transportation<br>• No diagnosis of a neurological disease<br>• Telephone Interview of Cognitive Status score > 25<br>• Cognitive adjudication decision of cognitively normal | • Current diagnosis of an Axis I or II disorder including Major Depression<br>• History of major psychiatric illness including schizophrenia (not including general anxiety disorder or depression (Geriatric Depression Scale [GDS] $\geq 9$))<br>• Current treatment for cancer – except non-melanoma skin cancer<br>• Neurological condition (MS, Parkinson's, Dementia) or brain injury (Stroke)<br>• Type I Diabetes, Insulin-dependent Type II Diabetes, uncontrolled Type II diabetes (defined as an HbA1c level > 10)<br>• Current alcohol or substance abuse or treatment for abuse in the past 5 years<br>• Current treatment for congestive heart failure, angina, uncontrolled arrhythmia, deep vein thrombosis (DVT) or another cardiovascular event<br>• Myocardial infarction, coronary artery bypass grafting, angioplasty or other cardiac condition in the past year<br>• Claustrophobia or inability to complete the MRI scan due to metal implants (pacemaker, stents) that are MR ineligible<br>• Color Blindness<br>• Engaging in >20 min of moderate intensity physical activity per day for 3 days or more per week<br>• Not local or able to travel 3 times per week to the exercise facility<br>• Travelling consecutively for 3 weeks or more during the study<br>• Unwillingness to be randomized to one of the three groups<br>• Current participation in an ongoing trial likely to influence exercise ability or cognitive function (e.g., mindfulness training). |

The University of Pittsburgh team (UPitt) used mass emails to a university operated registry of individuals interested in research. These mass emails targeted adults over the age of 65 years, or URM populations that lived in the greater Pittsburgh area. A similar approach was taken for commercially purchased mailing lists. Mailings to specific ZIP™ codes were made multiple times to increase familiarity with the trial. Efforts were made to emphasize ZIP™ codes with higher percentages of URM. Finally, the team regularly distributed brochures at local doctor offices, and purchased advertisements on public transportation, in arts and entertainment magazines and playbills, and in the local African American newspaper. UPitt employed 3 staff members (2 full time and 1 part time on this task) to complete phone screens and arrange study promotion activities.

Northeastern University in Boston, MA (NEU) sent postcards approximately every 3 weeks to commercially purchased mailing lists in ZIP™ codes with high URM populations, residents in the target age range, and high proportions of men. NEU employed one full and one part-time individuals to perform phone screening, along with a portion of the trial coordinator’s time.

The University of Kansas Medical Center team (KUMC) uses a centralized recruitment infrastructure which provides initial screening and triage of prospective participants for 10-30 studies at any time.[11] This reduces screening burden on the study team, but potentially increases recruitment personnel costs. Specific to the IGNITE Trial, the KUMC team performed two mass mailings, from two separate university-based participant registries to potential study candidates in the Kansas City metropolitan area, based on inclusion criteria. The team performed approximately 160 community talks during the recruitment period, including promotion of IGNITE and other studies. KUMC employed one full-time recruitment specialist and one-to-three part-time phone screeners for IGNITE.

Analyses of number of people screened and consented were performed on all available data. We assumed missing data was randomly occurring. We used Cochran-Mantel-Haenszel testing to account for site effects. We also performed random forest and classification tree analysis to explore demographic patterns related to participation. Due to low numbers of those screened or enrolled in some race categories, individuals identifying as non-White were collapsed into a single category for analysis. All analyses were performed with R v3.6.2.[19]

## RESULTS

Recruitment for IGNITE began on September 5, 2017, with a goal of randomizing 639 participants across three sites (Boston, Pittsburgh, Kansas City) by December 2020 with an equal number (n=213) at each site. We suspended recruitment and enrollment activity on March 13, 2020 at the direction of the UPitt trial coordinating center and local institutional guidance in response to the SARS-COV-2 pandemic. At the time of the suspension 2758 individuals had participated in screening and 487 had been randomized (76% of target). The study was on target for reaching randomization goals. Figure 1 shows the flow of participants through the study enrollment process.

**Figure 1.**
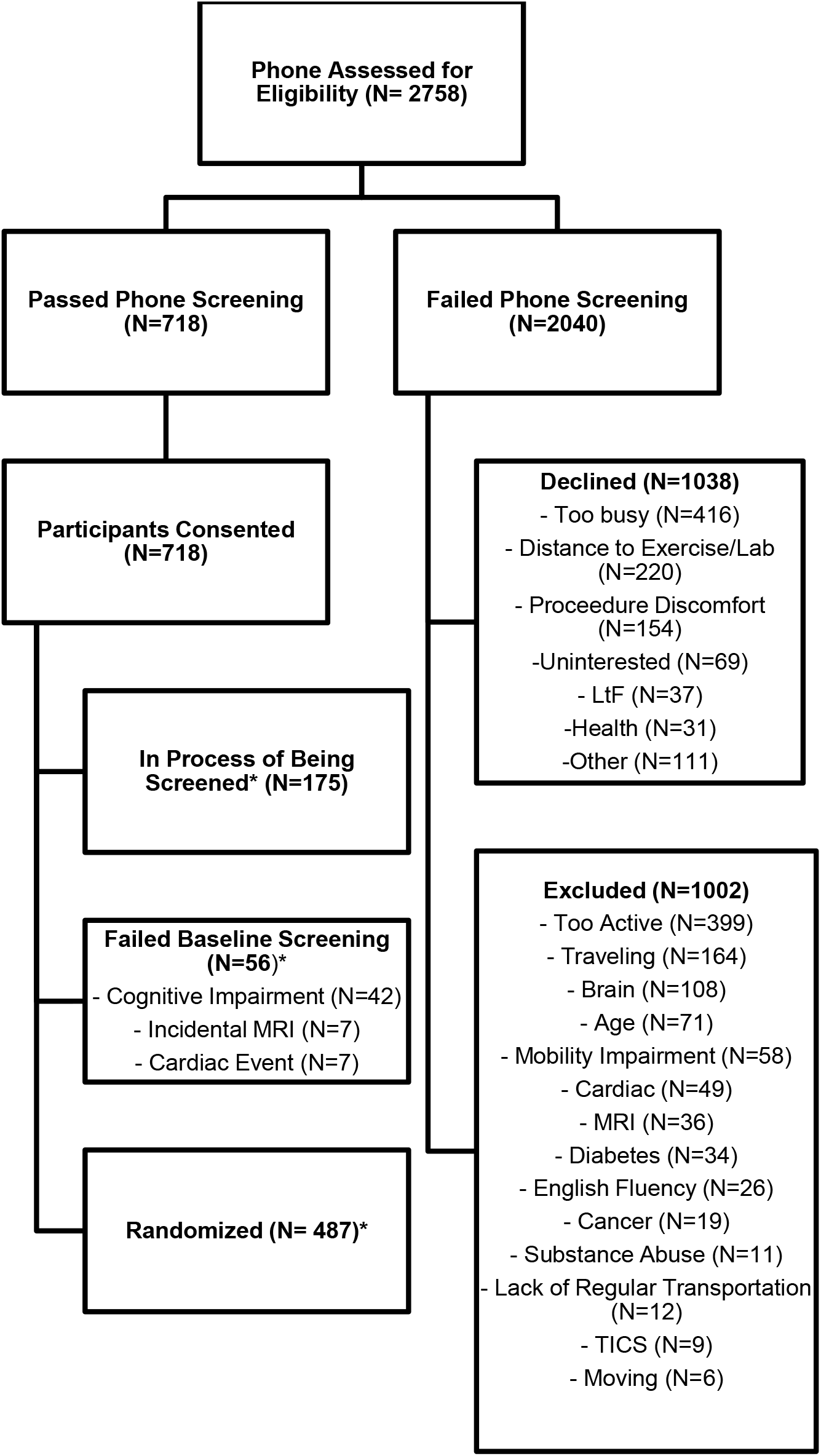
Screening and Enrollment Flow.

### Sources of Referral

There were differences in the referral source based on demographics accounting for site. Individuals identify as White responded less frequently to direct mailings and more frequently to word of mouth information across sites, especially if word of mouth came from a current or prior research participant (X^2^[10]=25.9, p=0.003). Individuals 75 and older were more likely to respond to direct mailings, and less likely to hear about the study through a website (X^2^[10]=22.8, p=0.01) Those who ultimately consented to participate were more frequently referred by a community presentation or through word of mouth (X^2^[10]=19.6, p=0.3). There were no differences in the referral source between men and women, or Hispanic ethnicity across sites (p>0.6). Anecdotally at KUMC, many participants who identified a non-professional (i.e., non-health care provider) word-of-mouth referral were often generated from staff presentations, either as primary or secondary (“friend of a friend”) referrals. Table 2 shows known referral sources by known demographic category.

**Table 2.** Study referral sources by demographics

|  | Community<br>Presentation | Direct<br>Mailing | Active<br>Ignite<br>Participant | Print<br>Advert | Prior<br>Research<br>Participant | Professional<br>Word of<br>Mouth | Non-Professional<br>Word of<br>Mouth | Social<br>Media | TV/<br>News | Website | Other |
| --- | --- | --- | --- | --- | --- | --- | --- | --- | --- | --- | --- |
| Male | 10 (1.9%) | 254<br>(49.5%) | 19 (3.7%) | 17 (3.3%) | 34 (6.6%) | 11 (2.1%) | 19 (3.7%) | 6 (1.2%) | 13<br>(2.5%) | 53<br>(10.3%) | 77 (15%) |
| Female | 29 (3%) | 469 (49%) | 51 (5.3%) | 39 (4.1%) | 64 (6.7%) | 18 (1.9%) | 38 (4%) | 24<br>(2.5%) | 35<br>(3.7%) | 85 (8.9%) | 106<br>(11.1%) |
| African American | 2 (0.9%) | 141<br>(66.2%) | 5 (2.3%) | 5 (2.3%) | 3 (1.4%) | 3 (1.4%) | 4 (1.9%) | 0 (0%) | 8 (3.8%) | 17 (8%) | 25<br>(11.7%) |
| Asian | 0 (0%) | 11<br>(61.1%) | 0 (0%) | 1 (5.6%) | 0 (0%) | 0 (0%) | 2 (11.1%) | 0 (0%) | 3<br>(16.7%) | 1 (5.6%) | 0 (0%) |
| White | 37 (3.2%) | 525 (45%) | 64 (5.5%) | 48 (4.1%) | 91 (7.8%) | 26 (2.2%) | 42 (3.6%) | 30<br>(2.6%) | 32<br>(2.7%) | 119<br>(10.2%) | 153<br>(13.1%) |
| American Indian/<br>Alaska Native | 0 (0%) | 2 (66.7%) | 0 (0%) | 0 (0%) | 0 (0%) | 0 (0%) | 0 (0%) | 0 (0%) | 0 (0%) | 0 (0%) | 1 (33.3%) |
| Native Hawaiian/<br>Pacific Islander | 0 (0%) | 1 (50%) | 0 (0%) | 1 (50%) | 0 (0%) | 0 (0%) | 0 (0%) | 0 (0%) | 0 (0%) | 0 (0%) | 0 (0%) |
| Refused | 0 (0%) | 23<br>(74.2%) | 1 (3.2%) | 0 (0%) | 2 (6.5%) | 0 (0%) | 2 (6.5%) | 0 (0%) | 1 (3.2%) | 0 (0%) | 2 (6.5%) |
| Biracial | 0 (0%) | 3 (100%) | 0 (0%) | 0 (0%) | 0 (0%) | 0 (0%) | 0 (0%) | 0 (0%) | 0 (0%) | 0 (0%) | 0 (0%) |
| Other | 0 (0%) | 7 (46.7%) | 0 (0%) | 0 (0%) | 2 (13.3%) | 0 (0%) | 5 (33.3%) | 0 (0%) | 0 (0%) | 1 (6.7%) | 0 (0%) |
| Hispanic/Latino | 2 (4.1%) | 31<br>(63.3%) | 1 (2%) | 1 (2%) | 3 (6.1%) | 1 (2%) | 3 (6.1%) | 0 (0%) | 0 (0%) | 2 (4.1%) | 5 (10.2%) |
| Not<br>Hispanic/Latino | 37 (2.6%) | 680<br>(48.5%) | 69 (4.9%) | 54 (3.9%) | 95 (6.8%) | 28 (2%) | 52 (3.7%) | 30<br>(2.1%) | 44<br>(3.1%) | 136<br>(9.7%) | 177<br>(12.6%) |
| Ultimately Not<br>Consented | 14 (2%) | 403<br>(57.1%) | 23 (3.3%) | 23 (3.3%) | 28 (4%) | 6 (0.8%) | 16 (2.3%) | 13<br>(1.8%) | 24<br>(3.4%) | 58 (8.2%) | 98<br>(13.9%) |
| Ultimately<br>Consented | 25 (3.1%) | 332<br>(41.8%) | 47 (5.9%) | 36 (4.5%) | 70 (8.8%) | 23 (2.9%) | 41 (5.2%) | 17<br>(2.1%) | 32 (4%) | 80<br>(10.1%) | 91<br>(11.5%) |

Referral sources of the 1452 individuals who recalled or will willing to report how they heard about the IGNITE study. Percentages are specific to each demographic sub-category. For example, 66.2% (141/198) of individuals who identified as African American and reported a referral source heard about the study from direct mailings.

Figure 2 shows the subgroups of participants identified through classification tree analysis. The subgroup of participants who reported a community presentation, current IGNITE participants, word-of-mouth, printed advertisements, prior research participants, social media, TV or news as the sources of referral showed the highest proportion to ultimately consent (70% consented). Among the participants whose sources of referrals were direct mailings, social media, website, or others, the subgroup of females younger than 75 years old shows the second highest proportion of giving their consent (56% consented) and the subgroup of males younger than 75 years old shows the second lowest proportion of giving their consent (47% consented). The subgroup of participants who were older than 75 years old shows the lowest proportion of giving their consent (32% consented). This exploratory classification tree shows consistent results with the important factors identified using random forests with 1,000 trees: age (mean decrease accuracy 52.17; mean decrease GINI index 16.15), sources of referral (27.44; 38.72) and gender (16.40;7.05).

**Figure 2.**
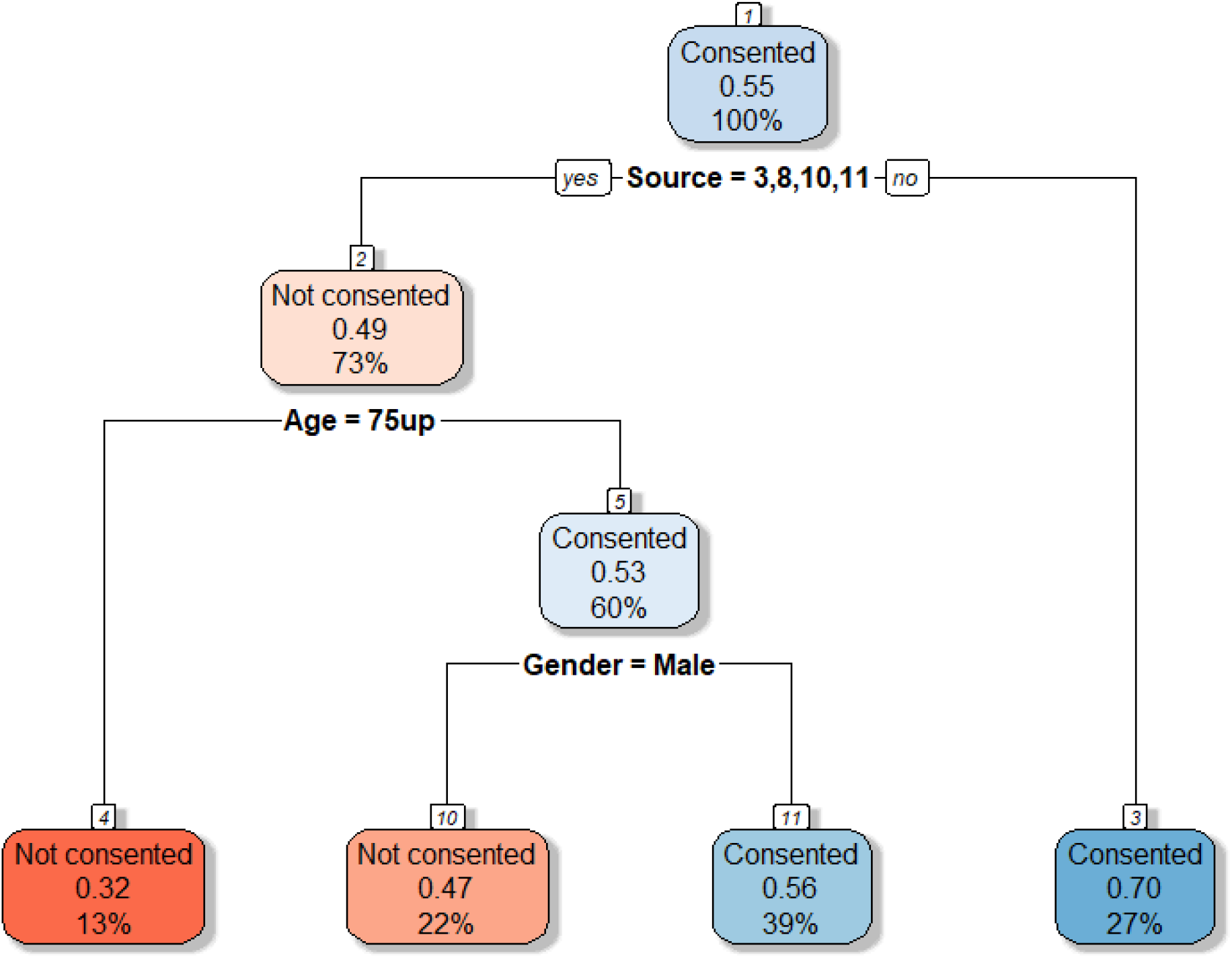
Subgroups of participants identified through classification tree analysis and the proportion of the consented participants by age (younger than 76 yrs old or not), gender, race (White or non-White), Ethnicity (Hispanic or non-Hispanic), and referral sources (1= Comm Presentation, 2=Current Ignite Pt, 3=Direct Mailing, 4=Non Professional Word of Mouth, 5=Print Ad, 6=Prior Research Participant 7=Professional Word of Mouth, 8=Social Media, 9=TV or News, 10=Website, 11=Other) using R package rpart. The minimum number of participants to split the node is set to 100 and complexity parameter is 0.01.

**Figure 3.**
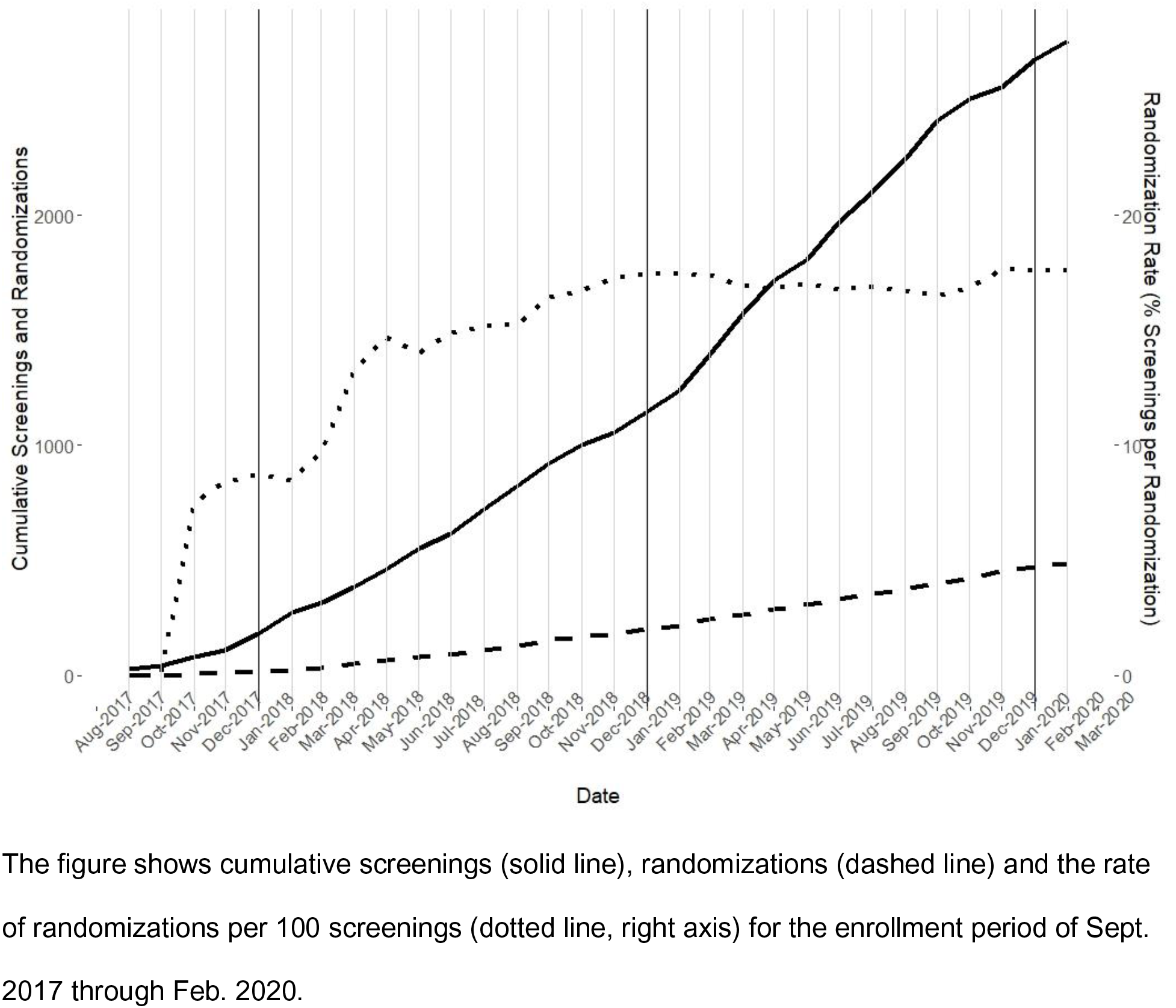
Cumulative Screenings and Randomizations During the IGNITE Enrollment Period

### Reasons for Dropout at Phone Screening

Of the 2758 individuals who began the screening process, 718 ultimately enrolled, and are discussed later. The remining 2040 individuals began the phone screening; 1038 (50.9%) declined to hear further information after the study design description. The most common reasons participants cited for declining were being “too busy” (n=416, 40.1%), distance to exercise facility/lab (n=220, 21.2%), and possible discomfort with procedures (n=154, 14.8%). The remaining 248 (23.9%) were not interested in the study premise, were lost-to-follow-up, or had another health condition or reasons they felt would limit their participation. There were no differences in reasons for lack of participation interest based on sex, race, age, or ethnicity (p>0.39). The other 1002 (49.1%) individuals completed the phone screen but were subsequently excluded based on entrance criteria. Reasons for exclusion are listed in Table 3. White-identifying individuals were more likely to be excluded for being too active and less likely to be excluded for insulin-dependent diabetes (X^2^[10]=35.6, p<0.001). Hispanic or Latino identifying individuals were more likely to be excluded due to heart disease and insulin-dependent diabetes, and less likely to be excluded due to ongoing reported physical activity, more than 60 minutes per week of moderate intensity physical activity (X^2^[10]=21.6, p=0.03). There were no differences in reasons for exclusion by gender or age (p>0.07).

**Table 3.**
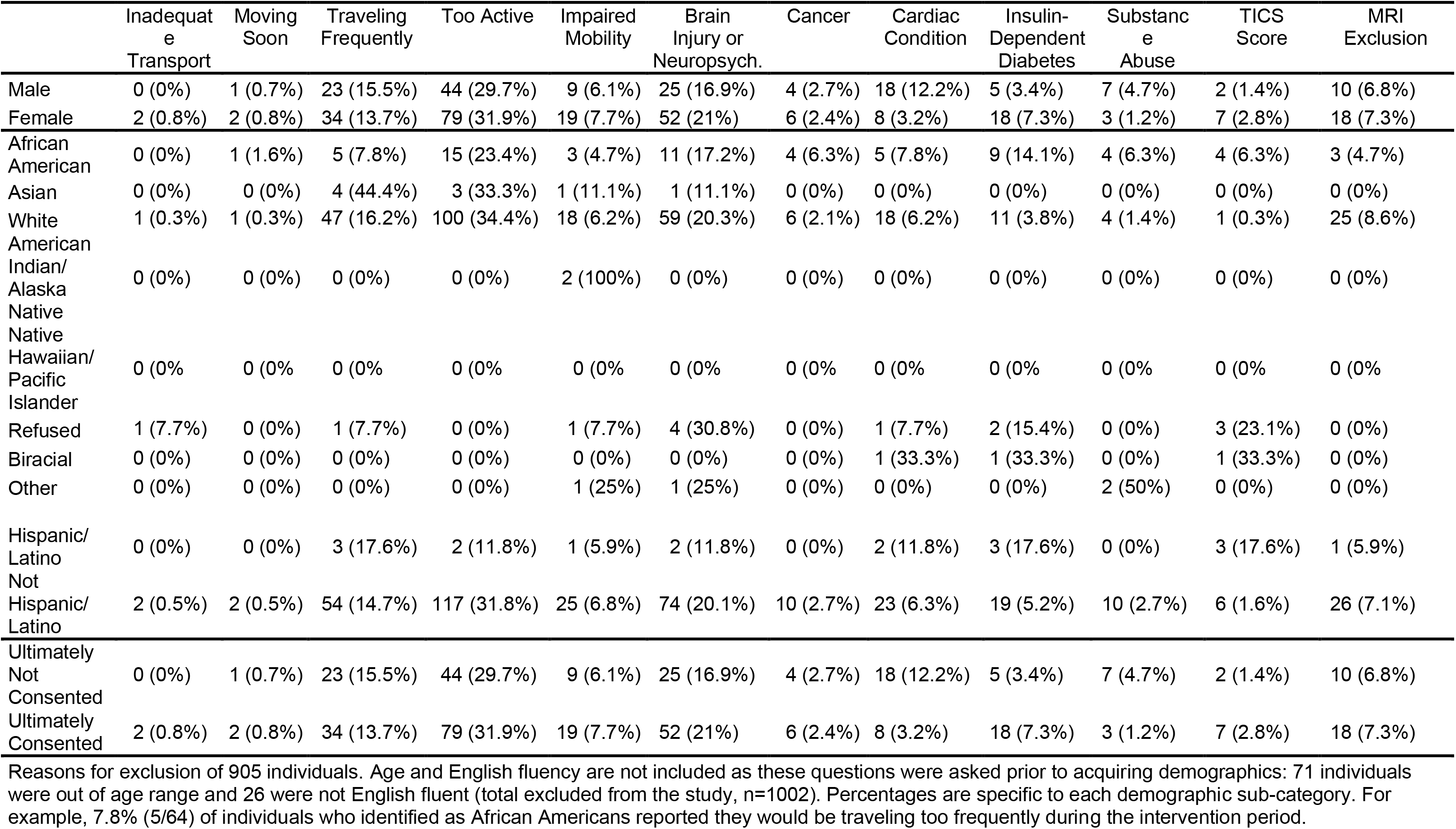
Reasons for exclusion by demographics.

### Baseline Screening

Of the remaining 718 individuals who cleared phone screening and consented to participate, 636 had completed baseline cognitive testing and been adjudicated by the neuropsychological team for normal cognition (the first step in baseline testing) at the time study procedures were halted. Forty-two individuals were judged by the neuropsychological team to have enough cognitive impairment to exclude them from further participation. Excluded individuals did not differ based on race, ethnicity or age, but were more likely to be male (9.6% vs 5.3%, X^2^[10]=4.2, p=0.04)

Once cognitive adjudication occurred, the remaining individuals were scheduled for a maximal exercise test or MRI according to participant and facility scheduling availability. There were no further demographic differences in exclusion from the study at this point (p>0.09).

Seven individuals were excluded due to an incidental MRI finding that was not subsequently cleared for continued participation. An additional seven individuals were excluded for poor performance or incidental cardiac findings during maximal exercise testing that were not subsequently cleared for continued participation.

### Randomized Participants

At the time study recruitment activities were paused in March 2020 due to SARS-CoV-2, the study had randomized 487 of the 718 consented individuals: 56 were excluded due to cognitive (n=42), MRI (n=7) or exercise test concerns (n=7) as noted previously. The remaining 175 were in various stages of screening or lost to follow-up. Table 4 shows the racial, ethnic and gender distributions of randomized individuals. Figure 2 shows the time series of screenings to randomizations. The ratio of randomized to screened individuals has stabilized at approximately 18:100.

**Table 4.** Demographics of randomized participants by site

|  | UPitt | KUMC | NEU | Total | Goal |
| --- | --- | --- | --- | --- | --- |
| Male | 54 (29.7%) | 49 (28.0%) | 36 (27.7%) | 139 (28.5%) | 195 (40.0%) |
| Female | 128 (70.3%) | 126 (72.0%) | 94 (72.3%) | 348 (71.5%) | 292 (60.0%) |
| African American | 28 (15.4%) | 7 (4.0%) | 26 (20.0%) | 61 (12.5%) | 99 (20.3%) |
| Asian | 1 (0.5%) | 2 (1.1%) | 4 (3.1%) | 7 (1.4%) | 29 (5.9%) |
| White | 151 (83.0%) | 161 (92.0%) | 94 (72.3%) | 406 (83.4%) | 359 (73.8%) |
| American Indian/<br>Alaska Native | 0 (0%) | 0 (0%) | 0 (0%) | 0 (0%) | 0 (0%) |
| Native Hawaiian/<br>Pacific Islander | 0 (0%) | 1 (0.6%) | 0 (0%) | 1 (0.2%) | 0 (0%) |
| Refused | 2 (1.1%) | 1 (0.6%) | 4 (3.1%) | 7 (1.4%) | 0 (0%) |
| Biracial | 0 (0%) | 0 (0%) | 0 (0%) | 0 (0%) | 0 (0%) |
| Other | 0 (0%) | 3 (1.7%) | 2 (1.5%) | 5 (1.0%) | 0 (0%) |
| Hispanic/Latino | 2 (1.1%) | 8 (4.6%) | 8 (6.2%) | 18 (3.7%) | 19 (19.0%) |
| Not Hispanic/Latino | 180 (98.9%) | 167 (95.4%) | 122 (93.8%) | 469 (96.3%) | 468 (81.0%) |

487 participants have been randomized to date. Percentages are specific to each site or the entire study.

### Recruitment Cost Results

Recruitment activities, staff and costs differed across sites. Total recruitment activity costs are summarized in Table 5. Promotional costs were derived from invoiced costs. Staff recruitment effort was based on estimated average weekly time spent in recruitment and promotional activities (e.g. materials and promotions coordination, speaking, participant phone screening). The total recruitment cost to date has been $276,596, with an average per-randomization cost of $590.43. Due to a variety of strategies employed, per-person randomization costs varied across sites, largely owing to different labor costs associated with recruitment; an estimated $199,187 were spent on staff time in recruitment. The estimated number of hours per randomization was 1.16, highlighting the need for dedicated recruitment staff in large-scale trials with high screening throughput.

**Table 5.** Recruitment costs and yield by site.

| Site | Promotional Cost | Staff Cost | Phone Screened | Number Randomized | Cost / Randomization | Adjusted Cost / Randomization* | Primary Strategy |
| --- | --- | --- | --- | --- | --- | --- | --- |
| UPitt | \$29319 | \$33541 | 1214 | 182 | \$345.38 | \$345.38 | Mailings, advertisement |
| NEU | \$35269 | \$68167 | 1080 | 130 | \$687.92 | \$687.92 | Mailings |
| KUMC | \$12816 | \$97479 | 464 | 175 | \$628.32 | \$628.32 | Mailings, presentations |
| Total | \$77404 | \$199,187 | 2758 | 487 | \$590.43 | \$553.87 | |

\*Per-person randomization costs adjusted by Sept. 2017 Consumer Price Index in each
metropolitan area using the 1967 base index for all items, relative to Pittsburgh.

## Discussion

Clinical trials require timely recruitment of a diverse group of participants for findings to be effectively generalized to the broader community. IGNITE has been designed as the definitive RCT of aerobic exercise on cognitive and brain health in older adults. Our interim analysis of recruitment and enrollment prior to the SARS-CoV-2 outbreak sought to identify potential patterns and biases that would decrease the generalizability of the findings and inform further enrollment. We found that direct advocacy approaches, such as a study team member making a community presentation or an individual advocating for participation provided the greatest number of individuals who met inclusion criteria and were ultimately randomized. We also found that exclusion criteria related to cardiometabolic disease precluded participation by those who are already typically underrepresented in research on aging.

Our analysis suggests that there are subtle but important process points at which individuals are more likely to be excluded. For example, medical history screening tended to exclude URM participants and men more frequently based on cardiovascular and metabolic disease, likely because of the known higher rates of cardiovascular disease in these populations. IGNITE’s comprehensive approach to cognitive testing also identified cognitive impairment in men more frequently than women. Prevalence of cognitive impairment is known to differ between sexes depending on etiology.[20] Our adjudication team did not assign etiology to the impairment, but the data suggest that exclusions based on a cognitive test battery may preferentially exclude older men more frequently than women.

Appropriate inclusion and exclusion criteria are critical for conducting safe and valid clinical research. However, these same criteria have the potential to exclude portions of the population that experience high comorbidity burden, introducing selection bias.[21, 22] It is unclear that IGNITE could safely loosen inclusion and exclusion criteria. However, given the higher rates of exclusion of men and URM, primarily African American and Hispanic individuals, a reassessment of methods and costs to allow individuals to safely exercise in the presence of comorbidity is likely warranted.

The IGNITE study team has worked to increase URM participation. However, not all sites have achieved high levels of URM representation. There are several possible reasons for this representation issue. Prior work has reported on the importance of long-standing relationships on the part of investigators and institutions to increase URM participation.[23] Each IGNITE site is in different stages of establishing relationships with URM communities. For example, the University of Kansas study team has established community partnership that are relatively new (<5 years) whereas the University of Pittsburgh study team has long-standing relationships and URM-dedicated RCTs ongoing. The Center for Cognitive and Brain Health at Northeastern was established two years ago and therefore is still building community partnerships. Approaches to expanding inclusion of underrepresented communities and men in research on aging and exercise need to be culturally tailored, responsive, and appropriate. There is no “one size fits all” approach. However, there is existing work that can serve as effective guides.[14, 24, 25] The IGNITE study team has begun sharing tested and vetted, culturally tailored education programs to increase engagement with African American and Hispanic communities.[26, 27]

Enrollment of men into trials is well recognized as a challenge.[28, 29] IGNITE expected and planned for higher numbers of women to enroll. However, enrollment of women exceeded predictions. There is some evidence that men are more likely to enroll in trials along with a female partner who also enrolls. There is also evidence that exercise may be viewed with skepticism or as a stereotypically feminine endeavor.[29] Going forward the IGNITE trial will consider soft restrictions on enrollment of White, non-Hispanic women unless enrolling accompanied by a male. The study team had previously planned for a small percentage of cohabitant enrollees but will consider relaxing this limit to increase male participants. In addition, teams will target community talks at traditionally male social organizations (e.g. VFW, Shriners, Elks).

The IGNITE study team expects the SARS-CoV-2 pandemic to significantly alter recruitment and study protocols. The team has begun revising and reducing in-person study visits to limit close contact, increase symptom screening, employ personal protective equipment for both participant and staff, and standardize SARS-CoV-2 appropriate cleaning procedures for all equipment and space. We anticipate that services and business necessary for clinical trial execution (public transit, community exercise facilities, etc.) will resume operations in an uneven manner across sites, further complicating study protocol. Contingency plans for at-home exercise and monitoring, and more frequent contact with study staff have been implemented. Standardized exercise videos have been distributed. Procedures for collecting data through electronic means (e.g. email, digital photos, REDCap surveys) have supplanted paper-based collection with IRB approval.

Anecdotally, sites are hearing a range of perspectives from current and prospective participants. Some individuals express enthusiasm for returning to in-person visits, community exercise, and the opportunity to participate. Others have expressed concern about the potential for infection at facilities that are closely affiliated with health systems treating patients with the COVID-19 acute respiratory syndrome that can result from SARS-CoV-2. The study team also anticipates elevated concern in URM communities regarding infection. URM individuals are experiencing greater rates of complicated infection and higher mortality rates in the US. While there is no definitive explanation for this disparity, there appear to be wide range of contributing structural and systemic inequities.[30-32] The study team anticipates additional and justified concerns about equitable and safe treatment in clinical trials that are compounded with the historical mistrust of clinical research in these communities.

Though the IGNITE recruitment experience can inform this and other exercise trials, several limitations should be considered. First, our phone screen flow did not capture all demographics and medical history on every potential participant. We do not know, for example, if the individual who did not want to have an MRI would have been excluded anyway for heart disease. This limits our ability to fully assess the impact of inclusion criteria on study diversity. We have made the assumption that individuals who were not interested in completing the phone screening were balanced in their race, ethnicity, and gender. This is in part due to Institutional Review Board requirements that the study be explained prior to collection of demographics or screening information. Second, the trial is not fully enrolled. It does not capture the potential effect of any increased investment in enrollment of specific populations, nor any damaging effect of SARS-CoV-2. We have listed our plans to support enrollment of men and individuals identifying as URM. The success of these initiatives remains to be quantified.

In conclusion, the IGNITE trial will provide essential information on the role of aerobic exercise in promoting sustained cognition and overall brain health as we age. The study team is making additional efforts to increase enrollment of men and URM to ensure the findings are generalizable to the broader American population of healthy older adults. Future studies in aging populations should consider process points that increase selection bias towards healthier White, non-Hispanic women.

## Data Availability

Data is available upon request from the principal investigator.

## Notes

### Competing Interest Statement

The authors have declared no competing interest.

### Clinical Trial

NCT02875301

### Funding Statement

R01 AG053952; R01 AG053952 04S1; P30 AG035982

